# Rapid rise in paediatric COVID-19 hospitalisations during the early stages of the Omicron wave, Tshwane District, South Africa

**DOI:** 10.1101/2021.12.21.21268108

**Authors:** Jeané Cloete, Annelet Kruger, Maureen Masha, Nicolette M du Plessis, Dini Mawela, Mphailele Tshukudu, Tabea Manyane, Lekwetji Komane, Marietjie Venter, Waasila Jassat, Ameena Goga, Ute Feucht

## Abstract

**Background:** South Africa reported a notable increase in COVID-19 cases from mid-November 2021 onwards, starting in Tshwane District, linked to rapid community spread of the Omicron variant. This coincided with a rapid rise in paediatric COVID-19-associated hospitalisations.

**Methods:** We synthesized data from five sources to describe the impact of Omicron on clinical manifestations and outcomes of hospitalized children (≤19 years) with positive SARS-CoV-2 tests in Tshwane District from 31 October to 11 December 2021, including: 1) COVID-19 line lists; 2) collated SARS-CoV-2 testing data; 3) SARS-CoV-2 genomic sequencing data; 4) COVID-19 hospitalisation surveillance; and 5) clinical data of public sector paediatric (≤13 years) COVID-19 hospitalisations.

**Findings:** During the six-week period 6,287 paediatric (≤19 years) COVID-19 cases were recorded in Tshwane District, of these 462 (7.2%) were hospitalized in 42 hospitals (18% of overall admissions). The number of paediatric cases was higher than in the prior 3 waves, uncharacteristically preceding adult hospitalisations. Of the 75 viral specimens sequenced from the district, 99% were Omicron. Detailed clinical information obtained from 139 of 183 (76%) admitted children (≤13 years; including all public sector hospitalisations) indicated that young children (0-4 years) were most affected (62%). Symptoms included fever (47%), cough (40%), vomiting (24%), difficulty breathing (23%), diarrhoea (20%) and convulsions (20%). Length of hospital stay was short (mean 3.2 days), and in 44% COVID-19 was the primary diagnosis. Most children received standard ward care (92%), with 31 (25%) receiving oxygen therapy. Seven children (6%) were ventilated; four children died, all related to complex underlying co-pathologies. All children and majority of parents for whom data were available were unvaccinated.

**Interpretation:** Rapid increases in paediatric COVID-19 cases and hospitalisations mirror high community transmission of SARS-CoV-2 (Omicron variant) in Tshwane District, South Africa. Continued monitoring is needed to understand the long-term impact of the Omicron variant on children.

**Research in context:** *Evidence before the study:* The announcement of the new Omicron (B.1.1.529) variant of the SARS-CoV-2 virus was made on 24 November 2021. Clinical characteristics, and disease profiles of children with COVID-19 before the arrival of Omicron have been described in the literature.

*Added value of the study:* This study describes the rapid rise in paediatric COVID-19-associated hospitalisations in Tshwane District in the Gauteng Province of South Africa – one of the first known epicentres of the new Omicron variant of the SARS-CoV-2 virus. The clinical picture as well as the steep increase in paediatric positivity rates and hospitalizations are described in detail from the perspective of a large South African health district, providing a broad overview on how the Omicron variant affects the paediatric population.

*Implication of all available evidence:* This study describes the clinical picture and outcomes in children in the current wave of SARS-CoV-2 Omicron variant infections by incorporating data from 42 hospitals at all levels of care in a large district within the South African health system. This provides novel paediatric data to assist global preparation for the impact of the Omicron variant in the paediatric setting.

## Introduction

On 24 November 2021 the Network for Genomic Surveillance in South Africa (NGS-SA; https://www.ngs-sa.org/ngs-sa_network_for_genomic_surveillance_south_africa/) announced a new variant of the SARS-CoV-2 virus, later named Omicron, first detected in a specimen collected on 9 November 2021. This announcement coincided with a significant rise in new SARS-CoV-2 infections in the Gauteng Province, heralding the start of South Africa’s fourth COVID-19 epidemic wave. The virus genome differed markedly from the Delta variant that had the highest prevalence during the third COVID-19 wave in South Africa. The Omicron viral genomic sequence includes 26-32 spike protein changes and 45-52 amino acid changes in total, causing worldwide concern regarding possible immune evasion. The World Health Organization (WHO) Technical Advisory Group on SARS-CoV-2 Evolution (TAG-VE) formally classified B.1.1.529 as the Omicron variant on 26 November 2021.^1–5^

Clinical characteristics and disease profiles of paediatric COVID-19 cases in South Africa in the first three waves have been broadly similar to descriptions in international literature, ranging from asymptomatic to mild-to-moderate disease in most children. In terms of clinically significant paediatric COVID-19 disease, the paediatric disease burden has not been severe during the first three COVID-19 waves, which occurred from June to September 2020 (first wave), November 2020 to February 2021 (second wave; Beta variant predominance) and June to October 2021 (third wave; Delta variant predominance).^6–20^

In marked contrast therefore were reports of the rapid rise in paediatric COVID-19 admissions, starting from mid-November 2021 onwards, in the Tshwane District, which is an urban district in the Gauteng Province of South Africa. The district has a population of 3,552,452 and a population density of 527 people/km^2^. There are nine public sector general hospitals serving the estimated 73% uninsured population, including two central, one academic, one regional and five district hospitals, together with four specialized hospitals. Additionally, the insured population has access to networks of private hospitals within the district.

This paper aims to describe the rapid rise in paediatric COVID-19 hospitalizations in 42 hospitals in the Tshwane District of South Africa during the start of the fourth COVID-19 wave, linked to the rapid spread of the Omicron variant in the geographic area, with a particular focus on the clinical manifestations and outcomes of hospitalized children.

## Methods

Data relevant to the Tshwane District were extracted from the following sources: 1) COVID-19 line lists for contact tracing activities (totals, with age break-downs); 2) SARS-CoV-2 testing data collated by the National Institute for Communicable Diseases (NICD); 3) SARS-CoV-2 genomic sequencing data from specimens obtained within the district through the Zoonotic Arbo and Respiratory Virus Research Group (ZARV), Department Medical Virology, National Health Laboratory Services (NHLS), University of Pretoria and NGS-SA from the global reference database for SARS-CoV-2 viral genomes (GISAID; www.gisaid.org); 4) COVID-19 hospitalisation data (DATCOV hospital surveillance system, collated by the NICD); 5) clinical data of public sector paediatric COVID-19 admissions (≤13 years), collected for the SA COVID Kids study and for local planning of paediatric clinical services.

Regarding the DATCOV surveillance system, 42 hospitals in Tshwane District are submitting hospitalisation data, including all public sector and 29 private sector hospitals (as on 11 December 2021). Due to restructuring during the COVID-19 pandemic one public sector district hospital was closely linked to its adjacent central hospital and subsequently counted as one hospital complex. The dataset includes information on patient numbers, age groups, gender, level of hospital care, length of stay and patient outcomes. Although the DATCOV system provides excellent overall disease surveillance, it lacks clinical detail as data entry is not clinician-driven. Therefore, clinical data from treating clinicians and hospital files were collated to supplement and verify the DATCOV data, including the clinical presentation, diagnoses, management, and outcomes.

For this study, paediatric COVID-19-related data were collated from 31 October 2021 to 11 December 2021 (epidemiologic weeks 44-49), to describe the period of rapid rise in paediatric hospitalisations during the initial phase of the forth COVID-19 wave. A laboratory-confirmed COVID-19 case was defined as any person who tested positive for SARS-CoV-2 on either real-time reverse transcription polymerase chain reaction (rRT-PCR) or antigen test conducted on samples obtained from nasopharyngeal or oropharyngeal swabs, with testing conducted at private and public sector laboratories. Access to SARS-CoV-2 testing was not markedly constrained - wider use of rapid antigen testing was the most important change in COVID-19 testing practices since the previous COVID-19 waves.

For genomic sequencing, clinical specimens were used from public sector clinics and hospitals in Tshwane District submitted to NHLS for SARS-CoV-2 rRT-PCR testing. Positive cases from 7 to 29 November 2021 (weeks 45-48) were collected by ZARV staff for genome sequencing. SARS-CoV-2-positive specimens with crossing point threshold values (ct) ≤30 were sent for Next Generation Sequencing to the Research Innovation & Sequencing Platform, University of KwaZulu-Natal (KRISP), as part of the NGS-SA initiative. Sequences were submitted to GISAID and assigned to lineages.

A COVID-19-associated admission was defined as any person who tested SARS-CoV-2 positive and was admitted to hospital, regardless of the reason for hospitalisation. Hospitals in the South African public health sector are divided into different levels of care, although for geographic reasons care is often first accessed for reasons of accessibility and not necessarily linked to disease severity. For descriptive purposes hospitals were grouped into central/academic and regional/district level, with high care and intensive care services available at the former, in addition to standard inpatient care available at all levels. Children who were referred were counted at the higher level, irrespective of the reason for referral.

The DATCOV national COVID-19 hospital surveillance system reports on children and adolescents ≤19 years of age. Descriptive statistics were used for demographic characteristics for tests, cases and hospitalisations in persons aged ≤19 years, and further stratified by age groups <1 year, 1-4 years, 5-9 years, 10-14 years, and 15-19 years, with adult population data shown for comparison. The crude admission rate was determined as the number of admissions in different age groups as a proportion of the population (as per Statistics South Africa mid-year population estimates for 2020), presented as admissions per 1,000,000 persons by age and week of admission. Paediatric hospital admissions in the South African public sector hospitals include children ≤13 years.

The Tshwane COVID-19 research study received permission from Ethics Committees of both Health Sciences Faculties in Tshwane (University of Pretoria Reference Number 822/2020; Sefako Makgatho Health Sciences University Reference Number SMUREC/M/54/2021:IR), together with the Tshwane District Research Committee. Additionally, the three large public sector hospitals are part of the SA COVID KIDS study (Reference Number EC048-11/2020; South African Medical Research Council). Ethics approval was also obtained for sequencing of COVID-19 samples (University of Pretoria Ethics Committee, Reference Number 101/2017).

## Results

The weekly test numbers, laboratory-confirmed COVID-19 cases, and COVID-19-associated admissions among residents in Gauteng Province, where Tshwane District is located, started rising rapidly from week 45 (7 November 2021), eight weeks after the end of the third wave (week 37). As of 11 December 2021, the district had recorded a cumulative number of 202,548 COVID-19 positive cases, with 36,152 admissions and 7,132 deaths, with the district’s four epidemic curves from March 2020 to date depicted graphically in Figure 1a.

**Figure 1:**
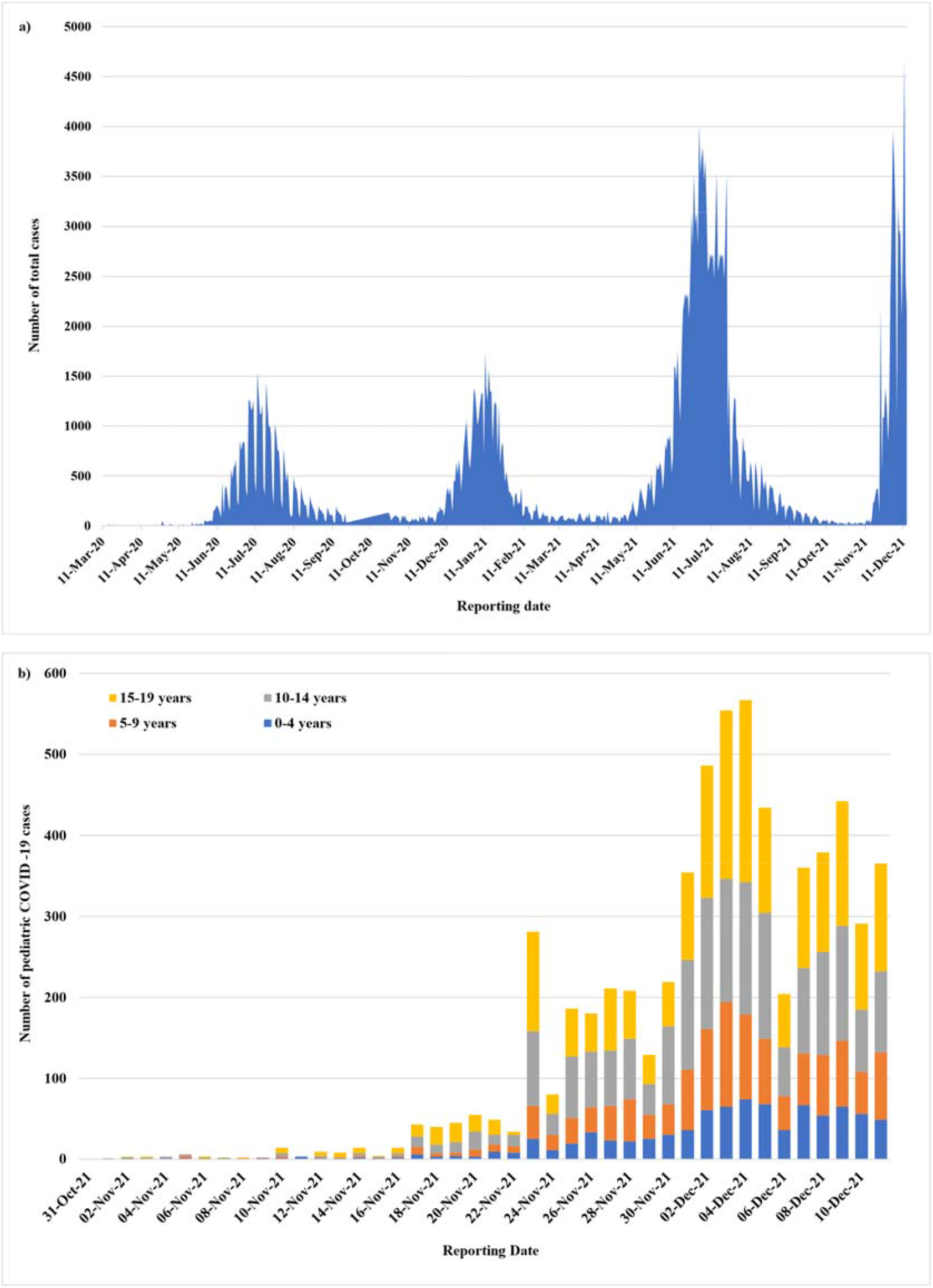
Daily COVID-19 cases in the Tshwane District (Tshwane District COVID-19 line list). a) Total daily COVID-19 cases (11 March 2020 - 11 December 2021); b) Number of paediatric COVID-19 cases per day per age category in the fourth epidemic wave (31 October 2021 - 11 December 2021)

In total 6,287 paediatric laboratory-confirmed COVID-19 cases were recorded on the Tshwane District COVID-19 line list between 31 October 2021 and 11 December 2021 (ages 0-4 years: 869; 5-9 years: 1,231; 10-14 years: 2,023; and 15-19 years: 2,164), Figure 1b.

The rapid rise in the number of SARS-CoV-2 testing numbers was studied in more detail using data from NHLS laboratories based at four large public sector hospitals in the district (Figure 2a). It shows high testing volumes, especially from the second wave onwards, also between the waves. During the waves the number of positive tests then increased sharply, as expected, together with the test positivity rate. The fourth wave is characterized by the steepest rise in percentage test positivity of all the waves, increasing very rapidly to above 40% (Figure 2b).

**Figure 2:**
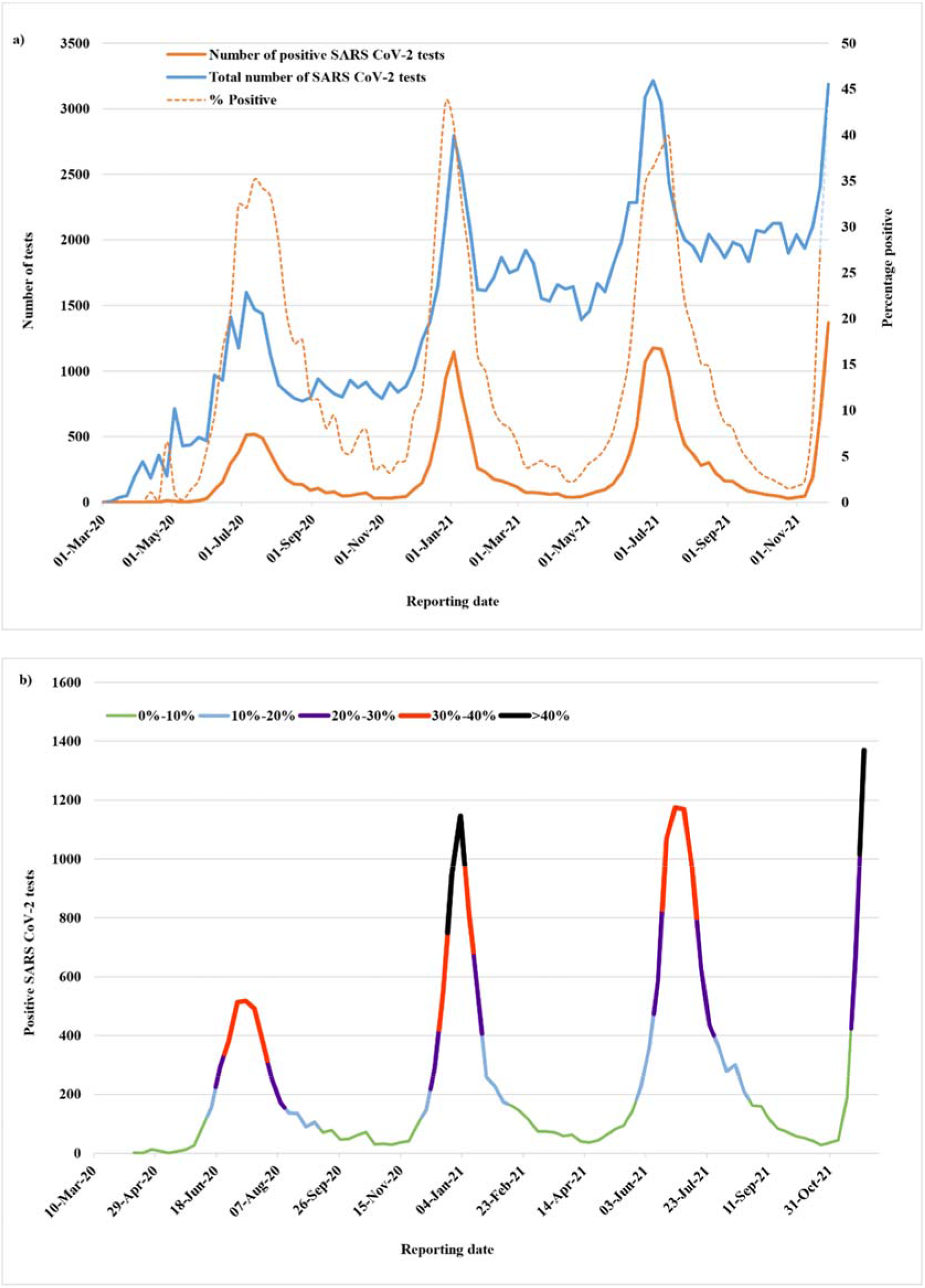
Total SARS CoV-2 testing numbers at four large public sector hospitals in Tshwane District, including tests conducted on adults and children. a) Total number of tests, number of positive tests and percentage of positive tests, and b) combined representation of test numbers and test positivity. (Laboratories located at three central/academic and one regional/district public sector hospital in the Tshwane District)

Genomic sequencing carried out on 75 SARS-CoV-2-positive specimens from public sector hospitals and clinics in Tshwane District between 7-29 November 2021 (weeks 45-48) were assigned as Omicron in 74 (98·7%) cases and Delta variant in one case only (1·3%). This suggests that the current increase in cases in the region is likely driven by the Omicron variant. The earliest detection of Omicron was on 12 November 2021. A more extensive analysis will be reported elsewhere.

COVID-19-associated hospitalisations started rising rapidly after the notable increase in cases in the district. Figure 3 depicts the hospitalisation numbers in all the four waves, as collected on the DATCOV hospital surveillance system for 42 hospitals in Tshwane. While overall hospitalisation numbers have to date been lower than in the previous three waves (Figure 3a), the hospitalisation numbers in children and adolescents (≤19 years) unexpectedly rose to higher levels than in the first three waves (Figure 3b). The overall number of COVID-19 admissions during the six-week period under review was 2,550, of which 462 (18%) were ≤19 years. The rise in paediatric COVID-19 admissions coincided with the rise in positive COVID-19 tests and test positivity, without any time delay (both occurred in week 46). This paediatric surge was seen in both the private and public sector hospitals (Figure 4c), with 181 admissions reported in the public sector and 281 in the private sector during the review period. The hospitalisations (overall and ≤19 years) showed a marked decrease in the last week of the study (week 49; 5-11 December 2021).

**Figure 3:**
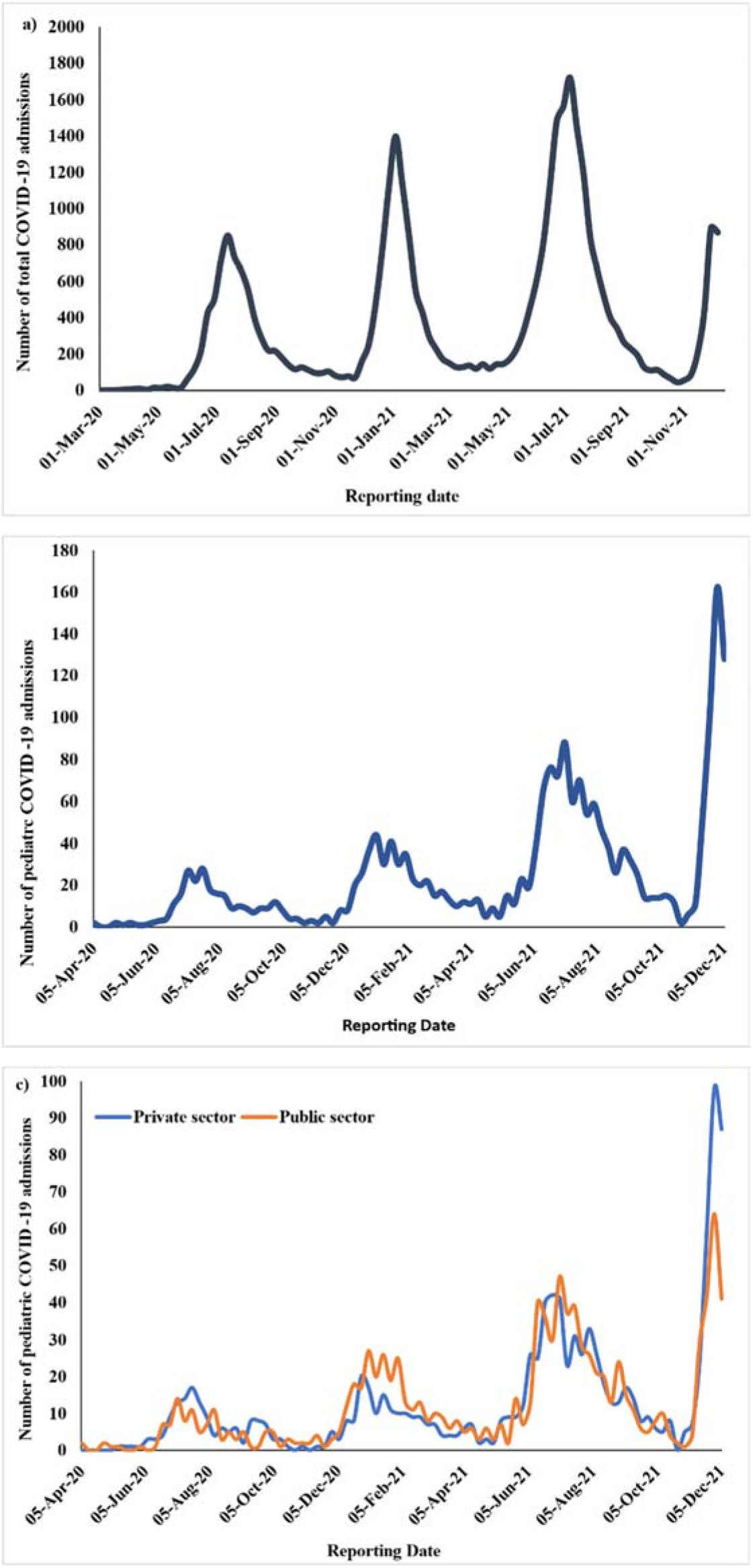
Number of hospital admissions in the Tshwane District: a) Total (adults and children); b) children and adolescents (≤19 years); c) children/ adolescents (≤19 years), as per public and private health care sectors (DATCOV; data from 42 hospitals) (1 March 2020 to 11 December 2021) ^23^

**Figure 4:**
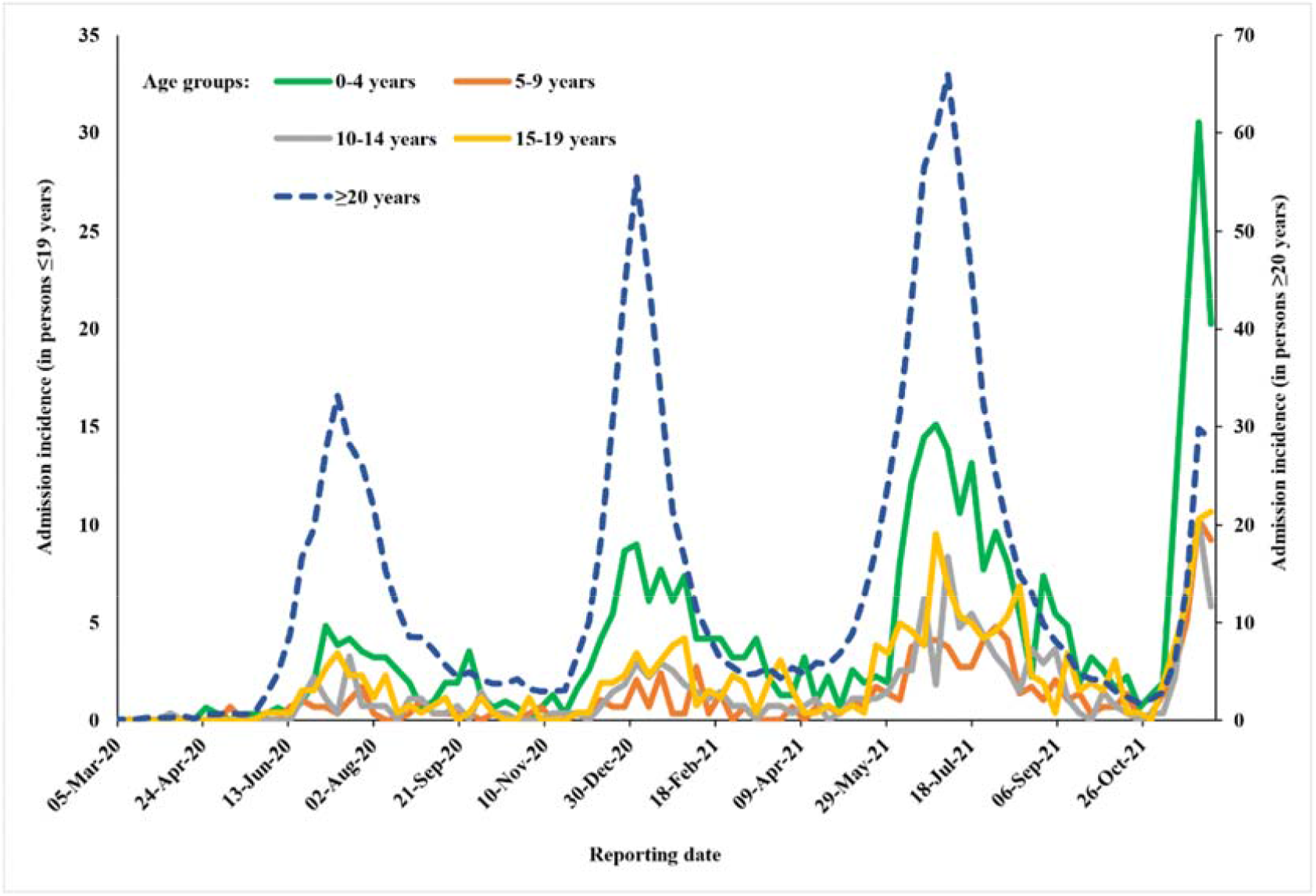
Admission incidence of COVID-19-associated hospitalisations in the Tshwane District, per age groups (DATCOV; data from 42 hospitals) (1 March 2020 to 11 December 2021)^23^

Further analysis of admission incidences, as shown in Figure 4, revealed that while in the first three waves paediatric admissions had lagged the adult admissions, this pattern was reversed in the fourth wave (particularly in the age group 0-4 years), confirming hospital-based clinicians’ observations of the unexpected rapid rise in paediatric COVID-19-associated admissions prior to rising adult hospitalisations.

Detailed clinical information on all COVID-19-associated hospitalisations in children ≤13 years, as recorded by local clinicians at all the public sector hospitals in the district, was analysed for the six-week period under review. Table 1, which is a summary of data from both DATCOV and the verified clinical information, shows the rapid increase in weekly hospitalisations from 14 November 2021 onwards, with numbers tapering off in the last week under review (week 49). This corroborated the trend already identified on the DATCOV data, indicating that the peak in COVID-19-associated paediatric hospitalisations had possibly been reached in the district.

**Table 1:** Admissions and deaths in COVID-19 positive children (≤13 years) in general public sector hospitals in Tshwane District (31 October 2021 to 11 December 2021) (Data sources: Verified clinical information; DATCOV)

|  |  | Week | Week 44 | Week 45 | Week 46 | Week 47 | Week 48 | Week 49 | Total |
| --- | --- | --- | --- | --- | --- | --- | --- | --- | --- |
|  |  | Date | 31 Oct – 6 Nov 2021 | 7 – 13 Nov 2021 | 14 – 20 Nov 2021 | 21 – 27 Nov 2021 | 28 Nov – 4 Dec 2021 | 5 – 11 Dec 2021 |  |
| **Admissions** | **Central/academic hospital 1** |  | 1 | 1 | 8 | 18 | 15 | 9 | **52** |
|  | **Central/academic hospital 2** |  | 0 | 1 | 3 | 5 | 11 | 13 | **33** |
|  | **Central/academic hospital 3** |  | 0 | 0 | 2 | 13 | 12 | 7 | **34** |
|  | **Regional/district hospital 1** |  | 1 | 1 | 0 | 5 | 3 | 5 | **15** |
|  | **Regional/district hospital 2** |  | 0 | 0 | 9 | 7 | 5 | 1 | **22** |
|  | **Regional/district hospital 3** |  | 2 | 0 | 0 | 3 | 6 | 0 | **11** |
|  | **Regional/district hospital 4** |  | 0 | 0 | 2 | 2 | 3 | 1 | **8** |
|  | **Regional/district hospital 5** |  | 0 | 0 | 0 | 2 | 1 | 5 | **8** |
|  | **Total** |  | **4** | **3** | **24** | **55** | **56** | **41** | **183** |
| **Deaths** | **Central/academic hospital 1** |  | 0 | 0 | 0 | 0 | 1 | 3 | **4** |
|  | **Total** |  | **0** | **0** | **0** | **0** | **1** | **3** | **4** |

Detailed clinical information is available for 139 (76%) of the 183 children hospitalized in all the public sector hospitals in the six-week period under review, including all children who died or had received high care or intensive care. The mean age of the hospitalized children was 4·2 years, ranging from newborn to 13 years, with most admissions in the <1 year age group (35%), and 62% of children in the 0-4 years category (Table 2). The male-to-female ratio was 1.28:1.

**Table 2:** Clinical features of 139 children (≤13 years) with COVID-19 who were admitted between 31 October 2021 and 11 December 2021 to public sector hospitals in the Tshwane District (clinically verified information)

| Age (in years) | Mean (SD) (range) | 4·2 (4·1) (0 – 13·5) |
| --- | --- | --- |
| **Age group** | <1 year | 48 (34·3%) |
|  | 1-4 years | 39 (28·1%) |
|  | 5-9 years | 34 (24·5%) |
|  | 10-14 years | 18 (12·9%) |
| **Gender** | Male | 78 (56·1%) |
|  | Female | 61 (43·9%) |
| **Presenting features\***<br>**(multiple presenting features possible)** | Fever | 58 (46·8%) |
|  | Cough | 50 (40·3%) |
|  | Vomiting | 30 (24·2%) |
|  | Shortness of breath/ Difficulty in breathing | 28 (22·5%) |
|  | Diarrhoea | 25 (20·2%) |
|  | Convulsions | 25 (20·2%) |
|  | Headache | 7 (5·6%) |
|  | Skin rash | 4 (3·2%) |
|  | Other (Body aches, painful joints, etc.) | 4 (3·2%) |
| **Duration of symptoms (days)** | Mean (SD) (range) | 1·7 (2·5) (1-14) |
| **Comorbidities\*\*** | None | 45 (39·5%) |
|  | Haematological/ oncological disease | 7 (6·1%) |
|  | HIV-exposed child | 6 (5·3%) |
|  | Diabetes mellitus type 1 | 6 (5·3%) |
|  | Cardiac disease | 4 (3·5%) |
|  | HIV-infected child | 4 (3·5%) |
|  | Cerebral palsy | 2 (1·8%) |
|  | Asthma | 1 (0·8%) |
|  | Various (including neonatal jaundice & sepsis, epilepsy, tuberculosis, neurosurgical conditions, burn wounds, other paediatric surgical and orthopaedic conditions) | 39 (34·2%) |
| **COVID-19 diagnosis in relation to hospital admission\*\*\*** | COVID-19 was primary clinical diagnosis | 58 (43·6%) |
|  | COVID-19 was a contributory diagnosis | 25 (18·8%) |
|  | Incidental COVID-19 diagnosis | 50 (37·6%) |
| **Laboratory results (on admission)\*\*\*\*** | Haemoglobin (g/dL) (mean; SD; range) | 12·5 (2·1) (8·3 -20·6) |
|  | Platelets (x 10<sup>9</sup>/L) (mean; SD; range) | 318 (122·6) (38-795) |
|  | White cell count (x 10<sup>9</sup>/L) (mean; SD; range) | 10·5 (5·3) (0·12-29) |
|  | Neutrophil count (x 10<sup>9</sup>/L) (mean; SD; range) | 5·6 (4·0) (0·03-20·4) |
|  | Lymphocyte count (x 10<sup>9</sup>/L) (mean; SD; range) | 3·8 (3·1) (0·06-16·4) |
|  | C-Reactive Protein (mg/L) (mean; SD; range) | 40·3 (82·4) (0-502) |
| **Length of stay† (days)** | Mean (SD) (range) | 3·2 (4·5) (0-30) |
| **Highest level of in-hospital care\*\*\*** | Standard ward care | 122 (91·7%) |
|  | Intensive care | 7 (5·3%) |
|  | High care | 4 (3·0%) |
| **Highest category of oxygen supplementation###** | None | 92 (74·8%) |
|  | Received any oxygen therapy | 31 (25·2%) |
|  | Nasal prong oxygen | 21 (17·1%) |
|  | High flow oxygen | 3 (2·4%) |
|  | Ventilation | 7 (5·7%) |
| **Outcome#** | Discharged | 102 (84·3%) |
|  | Still admitted (as on 15 December 2021) | 15 (12·4%) |
|  | Demised | 4 (3·3%) |
| **Final diagnosis ###** | Seizures | 25 (20·3%) |
|  | Acute gastroenteritis | 25 (20·3%) |
|  | Bronchopneumonia | 19 (15·4%) |
|  | Upper respiratory tract infection | 18 (14·6%) |
|  | Orthopaedic conditions | 8 (6·5%) |
|  | Sepsis | 7 (5·7%) |
|  | Diabetic ketoacidosis | 5 (4·1%) |
|  | Bronchiolitis | 3 (2·4%) |
|  | Croup | 3 (2·4%) |
|  | Appendicitis | 3 (2·4%) |
|  | Burns | 3 (2·4%) |
|  | Malnutrition | 3 (2·4%) |
|  | Near-drowning | 1 (0·8%) |

\*N=124; \*\*N=114; \*\*\*N=133; \*\*\*\*N=83; † N=115; # N=121; ### N=125 ### N=123

The clinical symptoms on presentation varied, with fever (47%) and cough (40%) noted as the two main symptoms, followed by vomiting (24%), difficulty in breathing (23%), diarrhoea (20%) and convulsions (20%). The mean length of stay was 3·2 days, with the longest stay of 30 days. Only four children were hospitalized for more than 14 days, all related to diagnoses other than COVID-19, including burn wounds, severe malnutrition, and tuberculosis. Just under 40% of admitted children had no underlying comorbidities, while in the others no single comorbid condition was found to be particularly common. The largest category was a combined group of unrelated conditions, which included neonates admitted with jaundice or sepsis; surgical patients (including orthopaedic, neurosurgical, and paediatric surgical conditions), burn wounds and epilepsy.

The most frequent clinical diagnoses linked to the hospitalisations were seizures and acute gastroenteritis (20% respectively), followed by respiratory infections namely upper respiratory infection and bronchopneumonia (15% each). The group who presented with uncomplicated seizures included six children who were outside the typical age range for simple febrile convulsions, as they were either less than one year or more than five years of age. All of them were discharged, and none had a diagnosis of bacterial meningitis.

COVID-19 was deemed by clinicians to have been the primary diagnosis in 43·6% of the hospitalized children, while in 18·8% it was deemed a contributory diagnosis and in 37·6% an incidental diagnosis. The last group included surgical patients with orthopaedic conditions, hydrocephalus, and appendicitis. No child was diagnosed with Multi-system Inflammatory Syndrome in Children (MIS-C), as per WHO case definition, in this six-week period.

Most children received standard ward care (92%), with 31 (25%) receiving oxygen therapy. Only three children (2%) received high flow oxygen therapy and seven children (6%) were ventilated. Reasons for ventilation included nosocomial sepsis, near-drowning, perforated appendicitis with septic shock, croup grade four and aspiration pneumonia, with one child ventilated for a presumed COVID-19 pneumonia. This child was an ex-premature baby with bronchopulmonary dysplasia and a new onset pneumonia, with other viral pathogens and pertussis excluded as possible causes.

The clinical presentations, diagnoses and management were similar at the central/academic and regional/district hospitals, apart from the few children (n=11) who required high care or intensive care. Seven children were referred to central/academic hospitals for higher level of care, including two COVID-19 related referrals and five referrals unrelated to COVID-19 disease. The great majority of children were discharged (84%), with 12% still in hospital at time of analysis.

There were four paediatric COVID-19-associated deaths recorded in this period, all occurring at one central/academic hospital with intensive care facilities. All children who demised presented with complex pathology and other significant diagnoses. The deaths were in children aged four months to ten years of age. Three of the children demised shortly after presentation due to their non-COVID-19-related reasons for hospitalization, and one child died of nosocomial sepsis. The in-hospital case fatality rate was 2·2% for COVID-19-associated admissions, but 0% if attributed to COVID-19 as primary diagnosis.

Currently in South Africa only children older than 12 years are eligible for COVID-19 vaccination and amongst the 121 children with data on vaccination status, none were vaccinated. Furthermore, of the 84 parents for whom COVID-19 vaccination data are available, 77 (92%) were unvaccinated and seven (8%) were partially or fully vaccinated.

## Discussion

We report a rapid rise in COVID-19 positivity and hospitalisations amongst children aged ≤19 years in Tshwane District, South Africa, linked with accelerated displacement of the Delta variant by Omicron, and high community transmission from mid-November 2021 onwards. The Omicron variant has been associated with lower antibody neutralization, higher infectivity, lower vaccine effectiveness and an increased risk for reinfection.^2,21^ The South African paediatric population is largely unvaccinated (vaccine eligibility started at age 12 years from October 2021). At the time of the start of the Omicron outbreak in Tshwane, 32% of adults in the district had received partial and 27% had received full COVID-19 vaccination. In the period under review, no boosters after the primary COVID-19 vaccination were being given in South Africa. Attempts were made to ascertain the COVID-19 vaccination status of the parents in the study, but this is unfortunately currently not well documented in clinical notes. Although many parents of the hospitalized children in this analysis were unvaccinated, they may have been partially protected from prior COVID-19 infection and by vaccinated adults interspersed within the adult population.^22^

The fourth COVID-19 wave started off from a low base, with evidence of very low levels of COVID-19 transmission in the communities of Tshwane District (low numbers of positive test results despite continued baseline testing, low test positivity rates, low COVID-19 hospitalisation numbers). The wave started earlier than expected, with paediatric hospital wards noticing marked increases in admissions of COVID-19 infected children from mid-November 2021 onwards, at much higher levels than in the previous three COVID-19 waves and uncharacteristically ahead of adult COVID-19-related admissions. The increased numbers of paediatric admissions, and rapid upward trajectory thereof, created logistical issues locally, as few COVID-19 paediatric hospital beds were available, as per experiences from the previous three COVID-19 waves. Coupled with the acute staff shortages due to COVID-19-related isolation and quarantine, this created a challenging environment in which to admit the unexpectedly large number of COVID-19 paediatric patients within the context of constrained resources.

Increases in positive COVID-19 tests and test positivity rates coincided with increased paediatric admissions (both occurred in week 46, with no time delay noted); this is in line with the high infectivity of Omicron. It is reassuring that 38% of COVID-19 diagnoses amongst hospitalized children were deemed incidental, with an additional 19% being a contributory diagnosis (not the primary diagnosis), indicating the rapid community-spread of the virus. National statistics from a private laboratory for November 2021 showed that the prevalence of most other seasonal respiratory viruses started to return to pre-pandemic levels in November 2021. Influenza A virus was the most prevalent virus in all four weeks of November 2021, followed by Rhinovirus (22%) and Adenovirus (12%), nationally, suggesting that Omicron is not displacing these viruses yet.^17^ We hypothesize that the high infectivity of Omicron with a short doubling time, high immune evasion, increasing vaccination coverage in adults, coupled with the fact that children wear masks less frequently than adults, could explain this unusual increase in children ahead of adults during the summer months in South Africa. COVID-19 lockdown regulations, including varying levels of intermittent school and preschool closures over the past 18 months, may also have affected exposure and natural immunity to common childhood illnesses amongst children.

The clinical picture of COVID-19 hospitalised children, as described in this paper, was varied and overlapping with other childhood illnesses, but the most significant finding was that 92% of children needed standard ward care with only seven (5%) needing intensive care and ventilation, 3% needed high care and 84% were discharged at the time of writing. The severity of disease was mostly mild-to-moderate, with a few children (25%) needing oxygen therapy and only one patient with comorbidities ventilated for COVID-19 pneumonia. The length of stay was also short (mean 3.2 days). Some previously unfamiliar clinical presentations were observed, including children presenting with convulsions, also in children with ages outside the norm for simple febrile convulsions, raising the possibility of an underlying encephalitis in these children presenting with COVID-19 infection. No current MIS-C cases have been recorded yet, although this may change as the fourth wave matures, with MIS-C known to be a late-onset, mostly post-COVID phenomenon. An analysis by the NICD from the pre-Omicron period (1 March 2020–28 August 2021) showed that of the 17,184 COVID-19-associated admissions among individuals aged ≤19 years (2·4%) were ventilated during the hospitalisation period. However, the hospitals included were mostly specialized, and by design admit sicker children. Furthermore, the NICD described an in-hospital case fatality risk (CFR) of 3·6% in the pre-Omicron era, slightly higher than the 2·2% in our study, with our analysis covering multiple public sector hospitals at all levels of care.^18^ Additionally of note is that in our study no child died primarily due to the COVID-19 infection.

Further research into the rapid spread of the Omicron variant will be crucial. Possible mechanisms for increased transmissibility of any airborne virus includes higher and/or longer viral shedding, change in tropism, better environmental survival, need for a lower infectious dose with more efficient viral entry, evasion of either innate or adaptive immune responses, and/or the ability to infect a new niche population, for instance younger children. All these possibilities will still need investigation to determine their relative contribution to the increased Omicron-related paediatric cases and hospitalisations, ahead of increased adult COVID-19-related hospitalisations. Of note, adult SARS-CoV-2 seropositivity from prior infection and/or COVID-19 vaccination was estimated at between 65-80% in South Africa in October 2021, with the lowest estimates in children 0-14 years (around 50%).^22^ When overall population immunity is low, then viral variants with high transmissibility will spread more easily, while in populations with higher immunity, the immune evasion properties become more important for facilitation of viral spread, with the latter being more likely in the current South African fourth wave with its rapidly rising COVID-19 cases.

Our analysis had several limitations: we used routinely collected datasets that have a lag in reporting and some incompleteness; additionally, we could not verify COVID-19 vaccination status - all were based on self-report, and this data element was not well captured in clinical notes. However, our paper has several strengths including that multiple data sources, including clinical data, were used to corroborate information, from a multi-disciplinary research team including clinicians, district-level health officials, epidemiologists and laboratory-based scientists.

Although our initial experiences in this fourth wave do not suggest that there is increased severity of COVID-19 disease amongst hospitalized children, infections and hospitalisations will be monitored in the district and updates provided. To date, the data have been reassuring.

## Data Availability

All data produced in the present study are available upon reasonable request to the authors

## Acknowledgements

Tshwane District management team; health care workers at public sector hospitals in Tshwane District (special mention: L Chumba, N Singh, M Maharaj, J Talma, E Sihlangu, T Muzinga, D Kutumela, J Mokwena, V Zulu); staff at Tshwane District Health Services responsible for COVID-19 surveillance (special mention: M van der Westhuizen, M Moshime-Shabangu); Tshwane District Clinical Specialist Team members (special mention: R Skhosana); DATCOV team (special mention: R Welch); SAMRC research staff (special mention: C Shabalala); laboratory team (special mention: Zoonotic arbo and Respiratory virus research group, Department of Medical Virology, University of Pretoria (M Davis; A Mendes; A Strydom); the National Health Laboratory Service Tshwane Academic division, Department Medical Virology, University of Pretoria, S Mayaphi and members of NGS-SA for sequencing information, https://www.ngs-sa.org/ngs-sa_network_for_genomic_surveillance_south_africa/).

